# Hospital-Based Donor Recruitment and Pre-Donation Serologic Testing for COVID-19 Convalescent Plasma

**DOI:** 10.1101/2020.07.20.20158048

**Authors:** Joanna Balcerek, Evelin Trejo, Kendall Levine, Paul Couey, Zoe V. Kornberg, Camille Rogine, Charlotte Young, Jonathan Li, Brian R. Shy, Jordan E. Taylor, Sara Bakhtary, Terence Friedlander, Kara L. Lynch, Caryn Bern, Jonathan H. Esensten

## Abstract

**Background:** Major blood centers perform serologic testing on potential COVID-19 convalescent plasma donors retrospectively after blood donation. A hospital-based recruitment program for COVID-19 convalescent plasma (CCP) donors may be an efficient way to prospectively identify potential donors.

**Study Design and Methods:** Patients who recovered from known or suspected COVID-19 were identified and recruited through medical record searches and public appeals. Participants were screened with a modified donor history questionnaire (DHQ), and if eligible, were consented and tested for SARS-CoV-2 antibodies (IgG and IgM). Participants who were positive for SARS-CoV-2 IgG were referred to a local blood center for convalescent plasma collection.

**Results:** Of 179 individuals screened, 128 completed serologic testing and 89 were referred for convalescent plasma donation to a local blood center (49.7% of those screened). IgG antibodies to SARS-CoV-2 were detected in 23/51 (45.1%) of participants with suspected COVID-19 and in 66/77 (85.7%) of participants with self-reported PCR-confirmed COVID-19. Testing was performed at a median of 38 days since last symptoms. Participant age positively correlated with anti-SARS-CoV-2 IgG and IgM levels. Time since last symptoms did not correlate with IgG or IgM levels. A wide range of SARS-CoV-2 IgG levels were observed.

**Conclusion:** A hospital based CCP donor recruitment program can prospectively identify potential CCP donors. Variability in SARS-CoV-2 IgG levels has implications for selection of CCP units for transfusion.

## Introduction

COVID-19 convalescent plasma (CCP) is being studied in multiple clinical trials to treat patients with COVID-19.^1–3^ However, recruitment of CCP donors and collection of CCP units initially lagged behind demand for this product.^4^ In normal times, blood centers try to exclude donors with specific infectious disease histories. For CCP donor recruitment by contrast, blood centers have the challenge of identifying donors with a history of confirmed SARS-CoV-2 infection. Hospitals can assist with the recruitment CCP donors by contacting recovered COVID-19 patients and referring them to blood centers.^5,6^

Per FDA guidance, COVID-19 convalescent plasma donors must meet all donor eligibility donor requirements for allogeneic blood donation. In addition, they must have evidence of prior infection with SARS-CoV-2 either by a positive PCR test at the time of illness, or a positive serologic test after recovery, if prior diagnostic testing was not performed. Donors must be asymptomatic for at least 14 days at the time of donation. In earlier versions of the FDA guidance, a nasopharyngeal swab that tested negative by PCR for SARS-CoV-2 and/or a 28 day symptom-free period was required before blood donation.^6^

In this study, we describe a two-center, hospital-based COVID-19 convalescent plasma donor recruitment program run in coordination with Vitalant. We screened study participants for eligibility for allogeneic blood donation and measured levels of anti-SARS-CoV-2 IgG and IgM antibodies. Participants who were positive for anti-SARS-CoV-2 IgG were referred for plasma donation. As part of this program, Vitalant agreed to send units from the first collection of referred CCP donors back to the referring hospital. These hospital-directed units are being used to support ongoing clinical trials.

## Materials and Methods

### Donor recruitment

Potential donors were recruited in April and May 2020 via medical record searches and public appeals. Medical records of PCR- or serology-confirmed COVID-19 patients at UCSF Health and Zuckerberg San Francisco General Hospital were identified, and where available, medical records were screened to exclude individuals who would not be eligible for allogeneic blood donation. Exclusion criteria included known disqualifying infections, medical conditions and medications, or continued hospitalization. Potential donors were then contacted by email and offered the opportunity to volunteer for the study. Recruitment materials for the study were made available to clinicians treating COVID-19 patients and to contact tracers at the San Francisco Department of Public Health. All potential donor information was recorded and stored exclusively in REDCap (v 9.5.25).

### Donor Screening

Participants were asked to provide the location, date, and method of testing for their COVID-19 diagnosis, if any, and answered a secure online version of a modified DHQ. Briefly, the modified DHQ consisted of yes/no questions developed by Vitalant with additional follow up questions requesting information (such as travel history) where appropriate. An automated scoring algorithm assigned participants to (1) donor eligible without follow up, (2) MD consult needed, or (3) donor ineligible. Clearly ineligible donors were screened out, while those reporting answers that required follow up were contacted by a study physician for clarification. Participants without a lab test confirmed history of COVID-19 were screened with the DHQ if they reported close contact with a known case and/or typical COVID-19 symptoms. Participants who passed the screen and who were judged likely to have been infected with SARS-CoV-2 were consented and referred for SARS-CoV-2 antibody testing.

### Donor Testing

Blood was collected by venipuncture and serum was tested for SARS-CoV-2 IgG and IgM using a Pylon 3D automated immunoassay system (ET Healthcare, Palo Alto, CA).^7^ This assay measures antibodies to the virus spike protein receptor binding domain (RBD) as described previously.^8^The assay result is expressed in relative fluorescence units (RFU). PCR testing for SARS-CoV-2 of nasopharyngeal swab samples was performed if a participant was 14-27 days after last symptoms. PCR was performed with the Abbott RealTime SARS-CoV-2 assay on an Abbott m2000 RealTime system.

### Analysis

Categorical variables were compared using the 2-tailed Chi square test. Continuous variables were compared using the Mann-Whitney test. Linear regression models were constructed for IgG and IgM levels versus days since last symptoms and age. All data analysis was performed using Prism (v8.4.2).

### IRB approval

This study received UCSF Institutional Review Board approval on 4/17/2020 (#20-30637).

## Results

An overview of the study process is provided in Figure 1A. Of 179 participants screened, 133 passed the screen, 128 were tested for SARS-CoV-2 antibodies, and 89 were referred for plasma donation. Among those screened, 44/179 (24.6%) of participants failed screening. Of participants who failed screening, 34 failed due to the DHQ and 10 had insufficient evidence of previous COVID-19. The most common reason for DHQ failure was travel history to areas with elevated risk of variant Creutzfeldt Jakob disease and men who reported recent sex with another man. Five participants who passed the screen did not set up a testing appointment.

**Figure legend:**
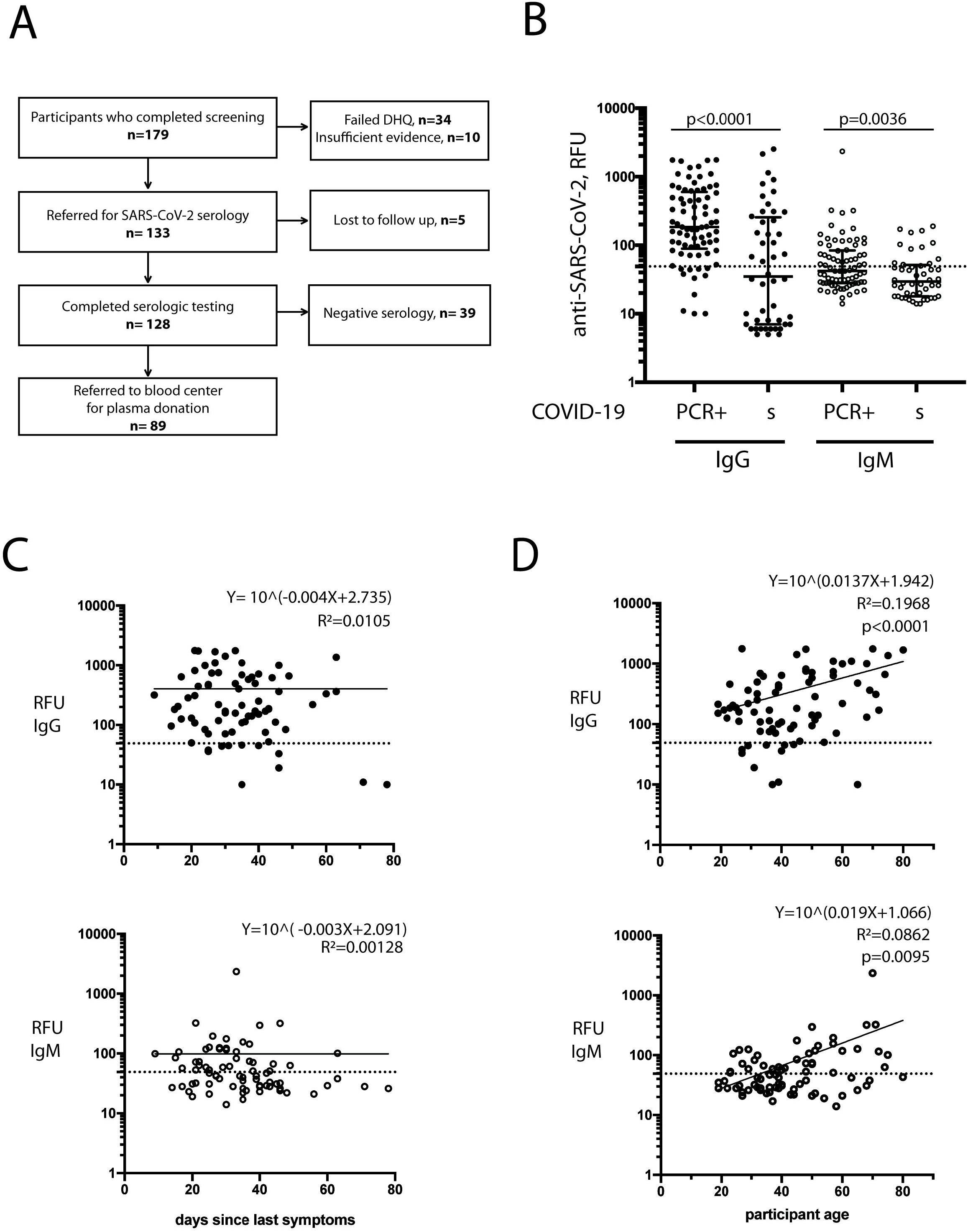
**(A)** Schematic of study process. **(B)** anti-SARS-CoV-2 IgG and IgM levels among participants with PCR-diagnosed (PCR+) COVID-19 (n=77), and suspected (indicated with “s”) COVID-19 based on history or serology (n=51). IgG and IgM levels are reported in relative fluorescent units (RFU). Dotted line represents RFU cutoff for seropositivity in this assay (50 RFU). (**C)** IgG and IgM level by days since last symptoms or (**D)** by age (in years) of participants among COVID-19 confirmed participants.

Characteristics of the study population who were tested are presented in Table 1. Participants were not referred for donation if they tested negative for anti-SARS-CoV-2 IgG. No significant differences were found between participants who were referred for SARS-CoV-2 plasma donation and those not referred when comparing sex, days since symptom resolution, participant age, or symptom severity. Among participants who were 14-27 days after last symptoms, 32 were tested for SARS-CoV-2 by PCR on nasopharyngeal swab specimens. Eight of 32 tested positive and were not referred for donation until they were 28 days after last symptoms.

**Table 1:**
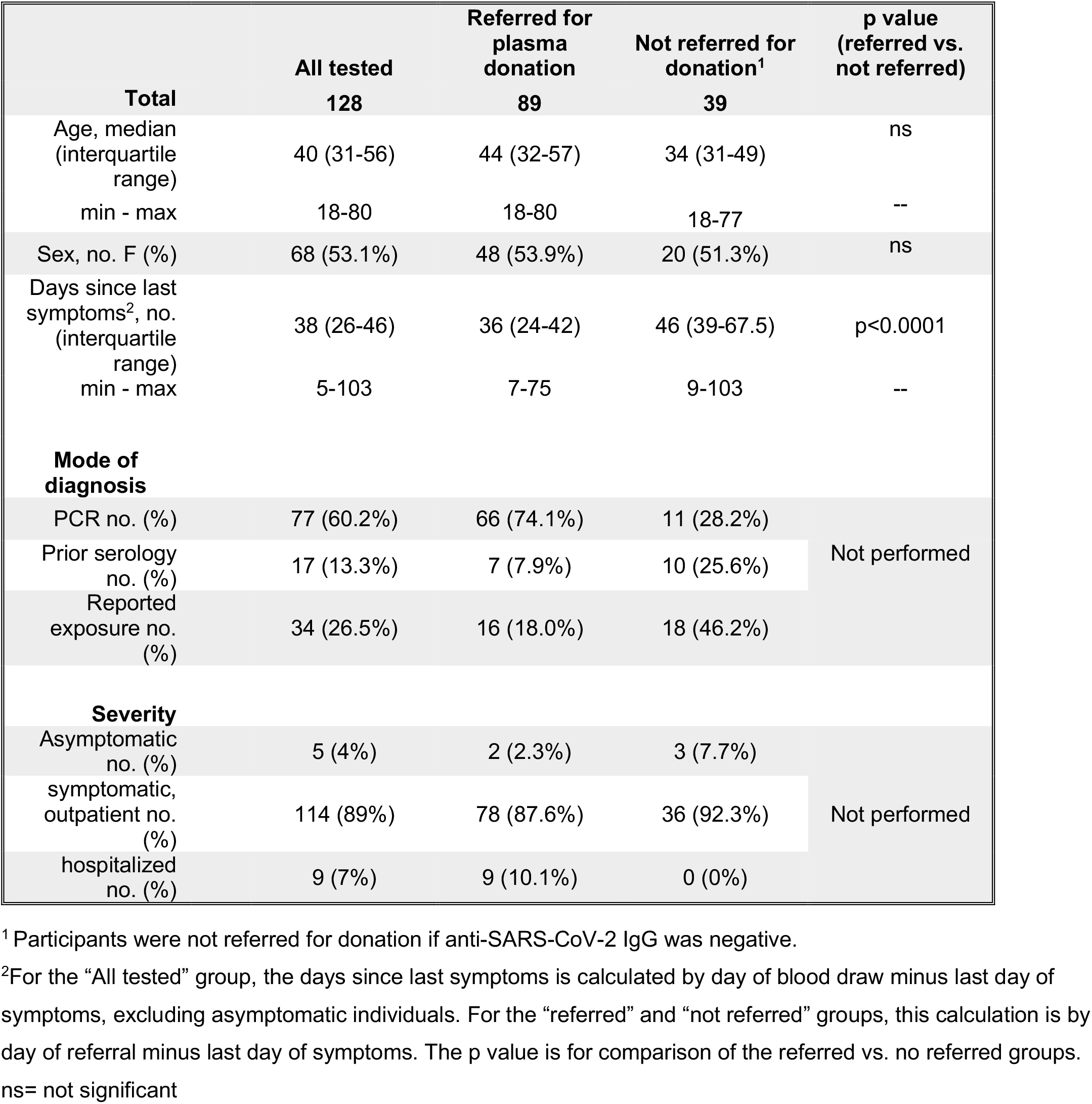
Participant demographics.

Anti-SARS-CoV-2 IgG was detected in 89/128 (69.5%) of tested individuals, including 66/77 (85.7%) of PCR confirmed and 23/51 (45.1%) of suspected cases (Table 2). IgM was detected in 48/128 (37.5%) of tested individuals: 35/77 (45.5%) of PCR confirmed and 13/51 (25.5%) of suspected cases (Fig 1B). Anti-SARS-CoV-2 IgG levels were significantly higher among PCR-confirmed participants (median 184 RFU, range 10-1764) compared to the COVID-19 suspected but PCR unconfirmed group (median 35 RFU, range 5-2520) (p<0.0001, Mann-Whitney test). There was no significant difference in IgG levels between PCR confirmed and unconfirmed participants if the values below the cutoff (50 RFU) were excluded in this comparison.

**Table 2:** Anti-SARS-CoV-2 serology results for all participants.

|  | **All** | **PCR +** | **suspected** | **p value** |
| --- | --- | --- | --- | --- |
| IgG positive (%) | 89/128 (69.5%) | 66/77 (85.7%) | 23/51 (45.1%) | p<0.0001<sup>1</sup> |
| IgM positive (%) | 48/128 (37.5%) | 35/77 (45.5%) | 13/51 (25.5%) | p=0.0224<sup>1</sup> |
| IgG RFU median (interquartile range) | 144 (37.5- 448) | 184 (89- 599) | 35 (7- 238.5) | p<0.0001<sup>2</sup> |
| min-max | 5- 2520 | 10- 1764 | 5- 2520 |  |
| IgM RFU median (interquartile range) | 38 (25.5- 71.25) | 42 (28- 84) | 29.5 (18- 50.5) | p=0.0024<sup>2</sup> |
| min-max | 14- 2334 | 14- 2334 | 14- 189 |  |

<sup>1</sup>By Chi square test for PCR+ vs suspected SARS-CoV-2 infection.
<sup>2</sup>By Mann-Whitney test for PCR+ vs suspected SARS-CoV-2 infection.

The cutoff value for a positive result in the serologic assay was set at 50 RFU for both IgM and IgG, which is 4 standard deviations above the mean of pre-COVID era plasma controls, and which resulted in 100% specificity during assay validation.^8^ IgG values were plotted against days since last symptoms among PCR-confirmed individuals (Fig 1C, top). We included only PCR confirmed individuals to minimize confounding from participants who may never have been infected. IgM level was plotted against days since last symptoms among individuals in the same group (Fig 1C, bottom). There was no correlation between anti-SARS-CoV-2 IgG or IgM levels and days since last illness.

Anti-SARS-CoV-2 IgG and IgM values were plotted against age of participants among PCR-confirmed cases (Fig 1D, top and bottom). A significant positive correlation was found between age and anti-SARS-CoV-2 IgG and IgM levels.

There was no significant difference in anti-SARS-CoV-2 IgG or IgM antibody levels between males and females. Participants who were hospitalized for COVID-19 had higher levels of anti-SARS-CoV-2 IgG than PCR-diagnosed non-hospitalized participants (median 742 vs. 171 RFU, p=0.029). There was no difference in IgM levels between these groups. However, this analysis is limited by the low number of previously hospitalized patients in this study (n=9).

## Discussion

Hospitals and public health authorities are uniquely positioned to aid in the recruitment of COVID-19 convalescent plasma donors through direct appeals to patients who have known positive test results. In this study, we pre-screened potential participants with a modified DHQ and a SARS-CoV-2 serologic test to maximize potential successful donation of CCP. In the first months of the COVID-19 pandemic, testing by PCR was limited to patients with the most severe symptoms. Therefore, we included participants in our study with exposure to known cases and/or typical symptoms of COVID-19 but who did not qualify to be tested. Rates of anti-SARS-CoV-2 seropositivity were higher in the PCR+ group than in the suspected but untested group (85.7% vs. 45.1% IgG positive). Given these results, potential donors with known exposure to a COVID-19 patient or with a history of typical symptoms but no prior testing should not be excluded from potential CCP donation. We did not test study participants for hemoglobin level, infectious disease markers, or anti-HLA antibodies. We have been informed by Vitalant that some of the referred donors from this study were deferred both at the donor center and after their CCP donation. Since many CCP donors are likely to be first-time blood donors or patients with comorbidities that predispose them to worse COVID-19 symptoms, deferrals may be more likely in this population.

An advantage of this study is that we directly contacted patients who had been diagnosed with COVID-19 to recruit them to donate plasma. We were able to access demographic and health information for COVID patients through hospital electronic medical records. However, as the number of different testing options increased, and test results were increasingly unlinked from any existing medical records, the task of identifying and contacting specific patients became more difficult. The San Francisco Department of Public Health, for example, allowed contact tracers to distribute study recruitment material but declined to give contact information of COVID-19 cases to our study team. Recruitment efforts for CCP donors targeting potential donors with a known COVID-19 diagnosis at the level of county or state health departments may be helpful to increase the donor pool. Although this study was done with IRB approval, approval to use patient health information for CCP donor recruitment may not be necessary per guidance from the US Department of Health and Human Services.^9^ Anti-SARS-CoV-2 IgG levels in 11 of the 77 PCR-confirmed cases fell below the assay cutoff, suggesting that one in seven infected individuals did not produce a serologic response. If a lower cutoff is used (30 RFU), an additional 8 PCR-confirmed cases would be considered IgG positive, resulting in an IgG positive rate of 74/77 (96%). Anti-SARS-CoV-2 IgG levels of 10 pre-COVID-19 plasma samples that we tested all fell below 10 RFU, which would argue in favor of counting the eight that fell above 30 but below 50 RFU into the “positive” category.

Given the observed variability in SARS-CoV-2 IgG levels in the participants in this study, a “positive” serology result may not translate into a clinically relevant dose of antibodies in a plasma unit. We have observed a up to a 50-fold difference in SARS-CoV-2 IgG levels between different CCP units that are confirmed positive for anti-SARS-CoV-2 IgG. This variability in anti-SARS-CoV-2 antibodies has been described by other groups but its cause is not clear.^10,11^ One limitation of this study is that we did not determine neutralizing antibody titers, anti-SARS-CoV-2 IgA levels, Ig subtypes, or antibody affinities. Other groups have shown correlation between antibody levels to the spike protein receptor binding domain and neutralization titer.^10,11^ Further work to characterize the functional heterogeneity of antibodies in different CCP donors is important to guide clinical use of CCP.

Due to the variability in anti-SARS-CoV-2 IgG levels, selection of specific units of CCP with higher antibody levels and neutralization titers may be required for clinical efficacy. Prospective testing of segments from CCP units in hospital blood bank inventories or labelling of units with antibody levels by blood suppliers could help with this selection. This approach would give patients a higher potential dose of antibodies with lower volume and donor exposure. However, requiring higher anti-SARS-CoV-2 antibody levels in CCP units may restrict CCP supply and may be costly for blood centers if many collected units fall below a designated cutoff.

Another consideration for using CCP is the amount of anti-SARS-CoV-2 antibody that a COVID-19 patient has already made. In 16 COVID19+ ICU patients at our institution we found anti-SARS-CoV-2 IgG levels at a median of 3595 RFU (range 2086 – 4009).^8^ These levels are far higher than levels measured in our CCP donors median 144 RFU (range 5-2520). Therefore, the clinical utility of transfusion of CCP in ICU patients with high baseline antibody levels must be carefully considered. This concern about the relative level of antibodies in the patient versus the CCP unit has led to suspension of one randomized controlled trial of CCP.^3^ Further work is needed to elucidate the optimal timing of treatment with convalescent plasma.

Finally, knowing the SARS-CoV-2 antibody status of a potential blood donor before blood donation has advantages. Donors can be scheduled for apheresis plasma collection rather than whole blood collection, which increases the amount of plasma collected and decreases the post-donation deferral period. Donors who are curious about their SARS-CoV-2 antibody status will have this information pre-donation and therefore will be more likely to be truthful on the DHQ at the time of donation. Finally, donors with higher levels of SARS-CoV-2 antibodies can be specifically recruited to ensure that only the most potent CCP products are distributed.

## Data Availability

The data that support the findings of this study are available on request from the corresponding author JE.

## Acknowledgements

The authors thank the study participants. We also thank Chav Doherty for technical assistance with study setup and execution.

